# Integrated Machine Learning-PanGWAS Reveals Chromosome-Encoded Persistence Networks and Plasmid Plasticity in Recurrent Urinary Tract Infection in *Escherichia coli*

**DOI:** 10.64898/2026.05.20.26353739

**Authors:** Sasikala R, Saisubramanian Nagarajan, Suma Mohan S

**Author notes:** Corresponding Author : Dr. Suma Mohan S.

## Abstract

**Background:** Recurrent urinary tract infections(rUTI) represent a major clinical challenge due to persistent clinical symptoms, repeated antibiotic exposure, and increased risk of multidrug resistance. Further clinical management of rUTI remains challenging, as existing diagnostic and treatment guidelines are largely designed for uncomplicated, acute infections. Though uropathogenic *Escherichia coli* (UPEC) is the predominant cause of community-acquired UTIs, pathogen-derived genomic features that may predispose certain *E. coli* strains to repeatedly establish infection are not fully understood.

**Methods:** To comprehensively dissect distinct genetic signals across genomic compartments that distinguish rUTI-associated isolates from those causing sporadic infection, the pangenome analysis in three different frameworks (i) Combined genomes (chromosome + plasmid), (ii) bacterial chromosomes only and (iii) plasmid-only was conducted. A comprehensive evaluation of population structure was performed using Gubbins, recombination-aware phylogeny IQTree, phylogroup distribution, pan-genome openness using Heap’s law, and plasmidome architecture using MOBSUITE.

**Findings:** Supervised machine learning models showed that the highest discriminatory performance was achieved using the combined genomic dataset (accuracy ∼0.98), and integration of feature-selected genes with PanGWAS (Pyseer and Scoary) identified a robust set of recurrence-associated genes, namely *cbtA, cbeA*, and *ldrD*, which were consistently detected across machine learning and association frameworks. Subsequent association rule mining further revealed cooperative gene networks enriched in rUTI isolates, particularly involving toxin-antitoxin modules and metabolic regulators.

**Interpretation:** Overall, this integrated ML-PanGWAS approach demonstrates that rUTI is a lineage-independent, polygenic phenotype encoded within a combined chromosomal-plasmid genomic context, providing new insights into the bacterial genomic architecture underlying recurrent disease and offering candidate biomarkers for future diagnostic and therapeutic development.

**Funding:** The Department of Biotechnology (DBT), Government of India (grant numbers: BT/PR40150/BTIS/137/81/2023 and the SASTRA TRR grant (SASTRA TRR SCBT OCT-23).

**Evidence before this study:** We searched PubMed, Scopus, and Web of Science for studies published from database inception to March 2026, without language restrictions, using combinations of the terms “recurrent urinary tract infection”, “uropathogenic *Escherichia coli*”, “pan-genome”, “genome-wide association”, “plasmid”, and “machine learning”. We included studies investigating genomic, phylogenetic, or functional differences between recurrent and sporadic UTI isolates. Previous studies have primarily used phylogenetic and single nucleotide polymorphism (SNP)-based approaches and reported limited genomic differentiation, with no consistent clustering or robust gene-level associations. Overall, existing evidence is heterogeneous and largely limited to single-layer genomic analyses.

**Added value of this study:** This study integrates pan-genome analysis across chromosomal, plasmid, and combined datasets with machine learning, genome-wide association analysis, and association rule mining. In contrast to previous SNP-based studies, we show that recurrent UTI isolates can be robustly discriminated only when chromosomal and plasmid features are analysed jointly. We identify reproducible recurrence-associated genes, including *cbtA, cbeA*, and *klcA*, and demonstrate that these genes form cooperative networks involving persistence, plasmid-mediated transfer, and metabolic adaptation, supporting a polygenic basis of recurrence.

**Implications of all the available evidence:** Our findings indicate that recurrent UTI is not driven by lineage alone but by coordinated accessory gene networks spanning chromosomal and plasmid compartments. Although certain sequence types are more frequently associated with recurrence, they are not exclusive and likely serve as backgrounds for enriched gene modules. These results highlight the importance of integrated genomic profiling for predicting recurrence risk and identifying persistence and adaptation pathways as potential targets for future diagnostic and therapeutic strategies.

## Introduction

Urinary tract infections (UTI) are among the most common bacterial infections worldwide and represent a significant threat to global health in both community and hospital-acquired infections. According to the Global Burden of Disease study, the number of UTI cases increased by 66.45% between 1990 and 2021, reaching an estimated 4.49 billion cases, with an age-standardized incidence rate (ASIR) of 5,531.88 per 100,000 population. The highest incidence of UTIs was recorded in women and older adult men (He et al., 2025). Recurrent urinary tract infections (rUTI) are defined as two or more episodes of UTI occurring within 6 months (Aggarwal & Lotfollahzadeh, 2022). Several risk factors, such as anatomical defects, incomplete antibiotic courses, inadequate fluid intake, lack of estrogen, increased antibiotic resistance, age, immunity, etc., could increase the risk of developing acute and recurrent UTIs in both men and women (Flores-Mireles et al., 2015). Recurrent urinary tract infections significantly affect the patient’s quality of life and health care costs due to repeated use of antibiotics, increased multiple drug resistance and persistent clinical complications (Hernández-Sánchez et al., 2025). Despite its prevalence, clinical management of rUTI remains challenging, as existing diagnostic and treatment guidelines are largely designed for uncomplicated, acute infections and provide limited guidance for recurrent or chronic presentations. Though rUTI is clinically classified as reinfection or persistence, the bacterial determinants that enable recurrence remain unclear. Uropathogenic *Escherichia coli* (UPEC) is the predominant pathogen among community-acquired UTIs, capable of colonizing the bladder and causing symptoms that range from mild cystitis to severe pyelonephritis. However, genomic features that may predispose certain *E. coli* strains to establish repeated infection are not fully understood. UPEC isolates exhibit a structured population in which a limited number of sequence types accounts for a large proportion of UTIs (Carter et al., 2023; García-García et al., 2025). Similarly, Maurizi et al., (2025) and Hidad et al., (2022) stated that rUTI isolates are enriched in certain sequence types. The genomic location of resistance genes can significantly influence bacterial phenotypes such as antibiotic susceptibility in UTI (Yang et al., 2021), suggesting that similar compartmental effects may contribute to persistence and recurrence in rUTI. Another study reported that plasmids carrying resistance genes can persist or vary across recurrent UTI episodes, indicating their potential role in adaptation and recurrence (Karami et al., 2021). Furthermore, it has been stated that recurrent UTI is driven by multiple bacterial mechanisms, including biofilm formation, persistence pathways, and virulence-associated genes, suggesting that recurrence is a multifactorial and potentially polygenic trait (Bhuiya et al., 2024). Based on these observations, in the current study, we tested the following hypotheses to systematically assess the genomic basis of recurrence across diverse *E. coli* isolates (i) Lineage hypothesis: rUTI isolates may belong to distinct phylogenetic lineages/clonal backgrounds as certain *E. coli* lineages are known to be associated with UTI (Nielsen et al., 2021) (ii) Genomic compartment hypothesis: Pathogen derived genetic patterns that distinguish rUTI from sporadic UTI isolates, may be encoded within chromosomal genes, plasmid-borne, or combined genomic contexts. (iii) Plasmidome hypothesis: rUTI-associated *E. coli* isolates differ from sporadic UTI isolates at the plasmidome level. (iv) Polygenic hypothesis: rUTI is associated with cooperative gene networks rather than single dominant virulence determinants. Guided by these hypotheses, the primary objective of this study was to identify population-scale genomic signatures that could distinguish rUTI-associated *E. coli* isolates from sporadic UTI isolates while acknowledging the host-immunity contribution to recurrence. To comprehensively capture distinct genetic signals across genomic compartments, the analysis was conducted in three parallel pan-genome frameworks: (i) Combined genomes (chromosome + plasmid), (ii) bacterial chromosomes only, and (iii) plasmid-only. This multi-layered approach enabled us to identify whether the discriminatory genetic patterns are encoded in chromosomal genes, plasmid-borne genes, or from their combined genomic context. In addition, this strategy allowed us to determine whether rUTI-associated isolates exhibit clonal or lineage-specific distinctions and whether recurrence is driven by single dominant factors or cooperative polygenic networks, thereby providing a comprehensive, hypothesis-driven framework to dissect the genomic determinants of rUTI.

## Materials and Methods

### Genome Selection

A total of 491 *E. coli* genomes associated with recurrent UTI and sporadic UTI isolates were compiled for this study using literature evidence and downloaded from the NCBI assembly database (https://www.ncbi.nlm.nih.gov/assembly/), and ENA (https://www.ebi.ac.uk/ena/). Downloaded datasets were classified into two phenotypic categories: rUTI group - recurrent UTI isolates and UTI group - sporadic UTI isolates based on clinical metadata (appendix 1). Based on the availability of sequencing data, assembled genomes were taken for the study only after passing the following threshold criteria: N50 ≥ 150,000 bp, Number of contigs < 100, Completeness > 99% using CheckM (v1.2.2)(Parks et al., 2015). Raw reads were subjected to initial quality assessment using FastQC (v0.12.1)(Andrews, 2010) and quality trimming was performed using trimmomatic (v0.39)(Bolger et al., 2014) and assembled using Unicycler (v0.5.0) (Wick et al., 2017) (de novo genome assembly). Only high-quality genomes were retained for pangenome and further downstream analysis.

### Pan-genome reconstruction

Genome assemblies were compartmentalized into chromosomal and plasmid components using MOB-suite(v3.1.9)(Robertson et al., 2018) followed by pan-genome analysis was performed using Panaroo (v1.3.4) (Tonkin-Hill et al., 2020) to evaluate the compartment-specific genomic signals through machine learning (ML). To minimise the impact of recombination on phylogenetic inference and ensure accurate lineage reconstruction, Gubbins (v3.3.0) (Croucher et al., 2015) was applied to the core genome alignment prior to maximum-likelihood tree construction using iQTree (v2.2.0) (Nguyen et al., 2015) and visualisation using iTOL(v6) (Letunic & Bork, 2021).

To assess genomic differences between rUTI and UTI isolates, population-level analyses were performed on the combined datasets. Population structure was further explored using principal component analysis (PCA), multidimensional scaling (MDS) based on gene-presence-absence matrices implemented in Python using the scikit-learn (Pedregosa et al., 2011) and matplotlib (Hunter, 2007) packages. Phylogroup classification was performed using Clermon Typing (EzClermont) (Beghain et al., 2018; Waters et al., 2020) for broad lineages grouping and Multilocus sequence typing (MLST) was performed according to the Achtman *E. coli* MLST (Wirth et al., 2006) scheme to assign sequence types (STs). Further pan-genome openness was evaluated by calculating Heap’s Law parameters using a custom python script, while core genome diversity and core genome shrinkage analysis were derived from Panaroo outputs. All figures were plotted using the Matplotlib Python packages (https://scipy.org). Plasmid sequences were extracted from the combined dataset and classified using MOB-suite (Robertson & Nash, 2018) for plasmidome analysis. Given the high variability of plasmid contigs, plasmid mobility, types of relaxases, mating pair formation types (MPF) and incompatibility groups (InC) were analysed to unravel whether rUTI exhibits a unique pattern against UTI isolates.

### Machine learning-based discrimination of rUTI and UTI

The generated two datasets, bacterial chromosome, plasmid was taken for further model performance along with the combined datasets to evaluate whether rUTI and UTI isolates could be discriminated based on specific genetic signatures. To identify significant genetic patterns, three complementary feature selection approaches were applied: Boruta (with cross-validation) (Kursa & Rudnicki, 2010), Chi square and Mutual information (Peng et al., 2005). A STRICT feature set was generated by taking the intersection of genes consistently selected across all three feature selection approaches (Chi-square, Mutual Information, and Boruta) and across cross-validation folds, representing high-confidence genetic signatures. In contrast, a RELAXED feature set was defined as the union of all genes identified by any of the three methods, capturing a broader set of potentially relevant features. These feature sets were generated independently for chromosomal, plasmid, and combined datasets and were used for downstream machine learning model training and evaluation. Multiple classifiers, including Logistic Regression, Random Forest, ExtraTrees, Support Vector Machine, and XGBoost (Chen & Guestrin, 2016), were evaluated. Based on the model performance, the combined gene-presence absence matrix was subsequently used for gene-level GWAS.

### Genome-Wide Association Analysis (Pan-GWAS)

To identify genes statistically associated with rUTI, Scoary (v1.6.16) (Brynildsrud et al., 2016) was used and to account for population structure correction kinship matrix based Pyseer (v1.3.11) (Lees et al., 2018) was performed on the combined datasets to identify genes associated with rUTI or UTI phenotypes. Significant genes were filtered using multiple testing correction, phenotype association strength and consistency across these three methods were considered as important genetic patterns that could distinguish rUTI isolates. Genes identified as significantly associated with rUTI only in any two of the methods (scoary vs pyseer, pyseer vs ML, scoary vs ML) were taken as seeded gene list and apriori algorithm was applied to the gene presence-absence matrices for that seeded gene list to identify co-occurring genes. This enabled detection of cooperative gene networks rather than single-gene effects. Final gene sets used from association rule was annotated post hoc to aid biological interpretation.

### External validation

To assess the robustness of the predictive models, an independent external dataset consisting 63 *E. coli* genomes was incorporated for validation (appendix 2). These genomes were not included in the model training phase and were obtained from independent sequencing projects available in the NCBI Assembly database and the European Nucleotide Archive (ENA). The external dataset comprised 31 recurrent UTI isolates and 32 sporadic UTI isolates, curated based on associated clinical metadata and literature evidence. Similar preprocessing and quality filtering criteria applied to the primary dataset were also applied to the external genomes to ensure consistency. Gene presence–absence matrices for these isolates were generated using the same pan-genome framework. Logistic Regression models trained on the combined genome feature space were evaluated on the external dataset to assess performance robustness. Performance metrics including accuracy, precision, recall, F1-score, and area under the ROC curve (AUC) were calculated to determine the robustness of the predictive signatures across independent cohorts. Strain-associated metadata along with the literature evidence for phenotypes, are available in appendix 1 and 2. A schematic overview of the analytical workflow, including genome processing, pan-genome reconstruction, machine learning, genome-wide association analysis, and external validation, is provided in appendix 3 pp 1.

## Results

### Recurrent UTI and sporadic UTI share an open pan-genome and similar genetic diversity

Pangenome analysis of 491 *E. coli* isolates identified a total of 22681 genes including 2070 core genes, 1292 soft-core genes, 2714 shell genes, and 16605 cloud genes. A very large cloud and shell genes represents the high genomic diversity of *E. coli* isolates and pan-genome openness. To further investigate the genomic compartmentalization of all the *E. coli* isolates taken for the study, pan-genome analyses were performed (i.e.) for chromosomes and plasmid assemblies separately. The chromosomal pangenome comprised 19,570 gene clusters, including 3,087 core genes, 269 soft-core genes, 2,492 shell genes, and 13,722 cloud genes. In contrast, the plasmid pangenome (468 plasmid-bearing isolates) consisted of 5,925 gene clusters, with no detectable core or soft-core genes and was dominated by 5,671 cloud genes and 254 shell genes (Supplementary Figure S2). The absence of conserved plasmid core genes and the predominance of cloud genes highlight extensive plasmid heterogeneity across isolates, whereas the chromosome retained a stable conserved genomic backbone. Heap’s Law modelling of pan-genome openness revealed that both rUTI and sporadic UTI isolates share similar open-pan genome architecture (rUTI = 0.759 and UTI=0.73) indicating gene acquisition within the population and support continuous horizontal gene transfer and genomic plasticity (figure1A). Comparative core genome analysis (rUTI = 0.016879; UTI = 0.014276) and genetic shrinkage analysis (Wilcoxon p-value =0.5051) revealed that both rUTI and UTI isolates exhibit similar core genome nucleotide diversity and no significant difference in genetic shrinkage (figure 1B and figure1C).

**Fig 1.**
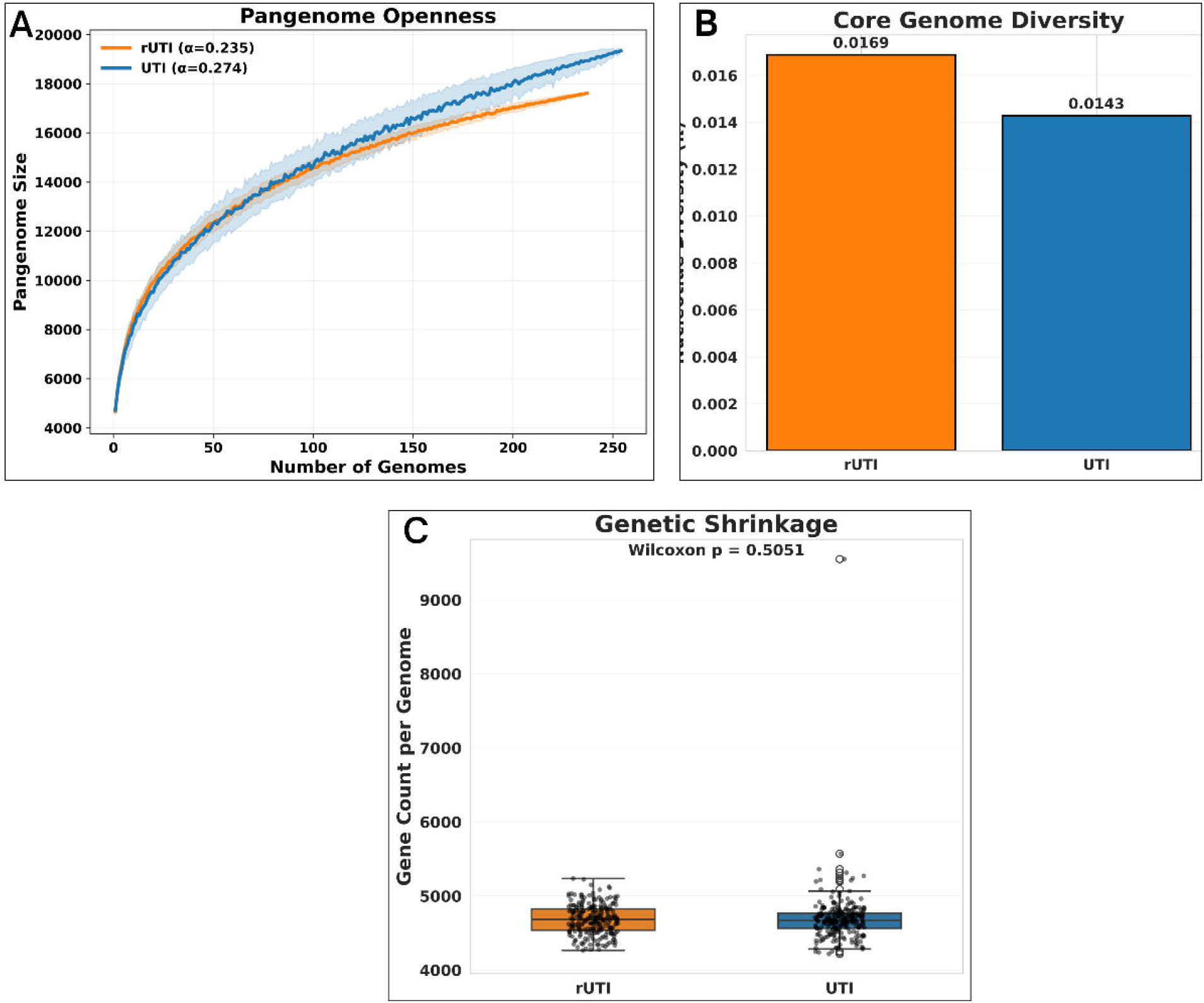

From the recombination-aware phylogenetic reconstruction of the core genomes, clustering analysis such as Principal Component Analysis (PCA) and Multi-Dimensional Scaling (MDS) using pan-genome gene-presence or absence matrix also revealed that there are no distinct clades or clusters observed across both rUTI and UTI (figure 2A and figure 2B).

**Fig 2.**
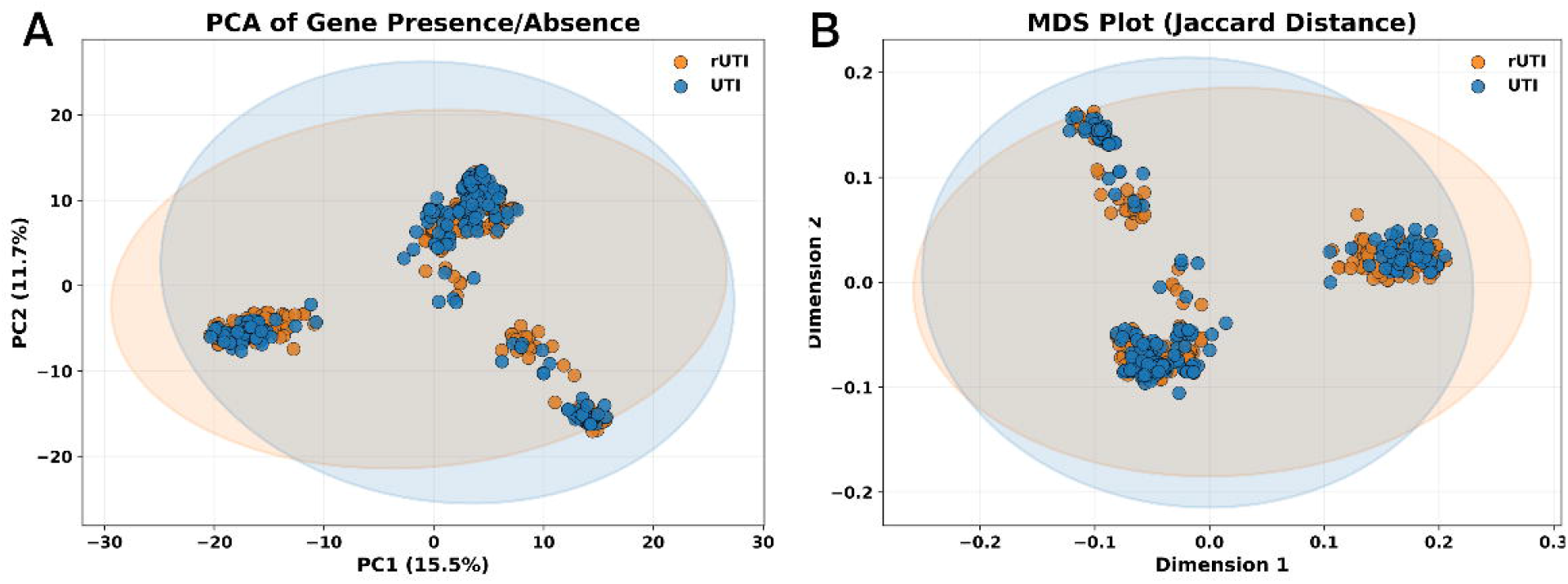

Further for understanding the population structure of the rUTI and UTI isolates, phylogroup classification and multilocus sequence typing (MLST) were performed. Phylogroups classification revealed that both rUTI and UTI isolates were predominantly distributed within phylogroup B2, which is consistent with the known predominance of extraintestinal pathogenic *E. coli* (ExPEC) lineages in UTI (appendix 4). However, differences in the relative prevalence of phylogroups such as D (rUTI-12.7%; UTI -7.5%) and B1 (rUTI-11%; UTI -6.8%) were observed between the two groups (figure 3A). To further resolve the lineage composition, MLST analysis was performed using the Atchman scheme (appendix 4). Several well-established uropathogenic lineages were identified across both groups, with ST131 emerging as the most prevalent sequence type. Interestingly, certain sequence types appeared preferentially enriched in rUTI isolates, including ST69, ST58, and ST404, which were either absent or minimally represented in sporadic infections (figure 3B). Conversely, some lineages such as ST1193, ST70, ST420, and ST12 were observed exclusively within the sporadic UTI group in this dataset. These findings indicate that while several globally disseminated ExPEC lineages contribute to both infection types, specific sequence types may exhibit differential association with recurrent disease.

**Fig 3.**
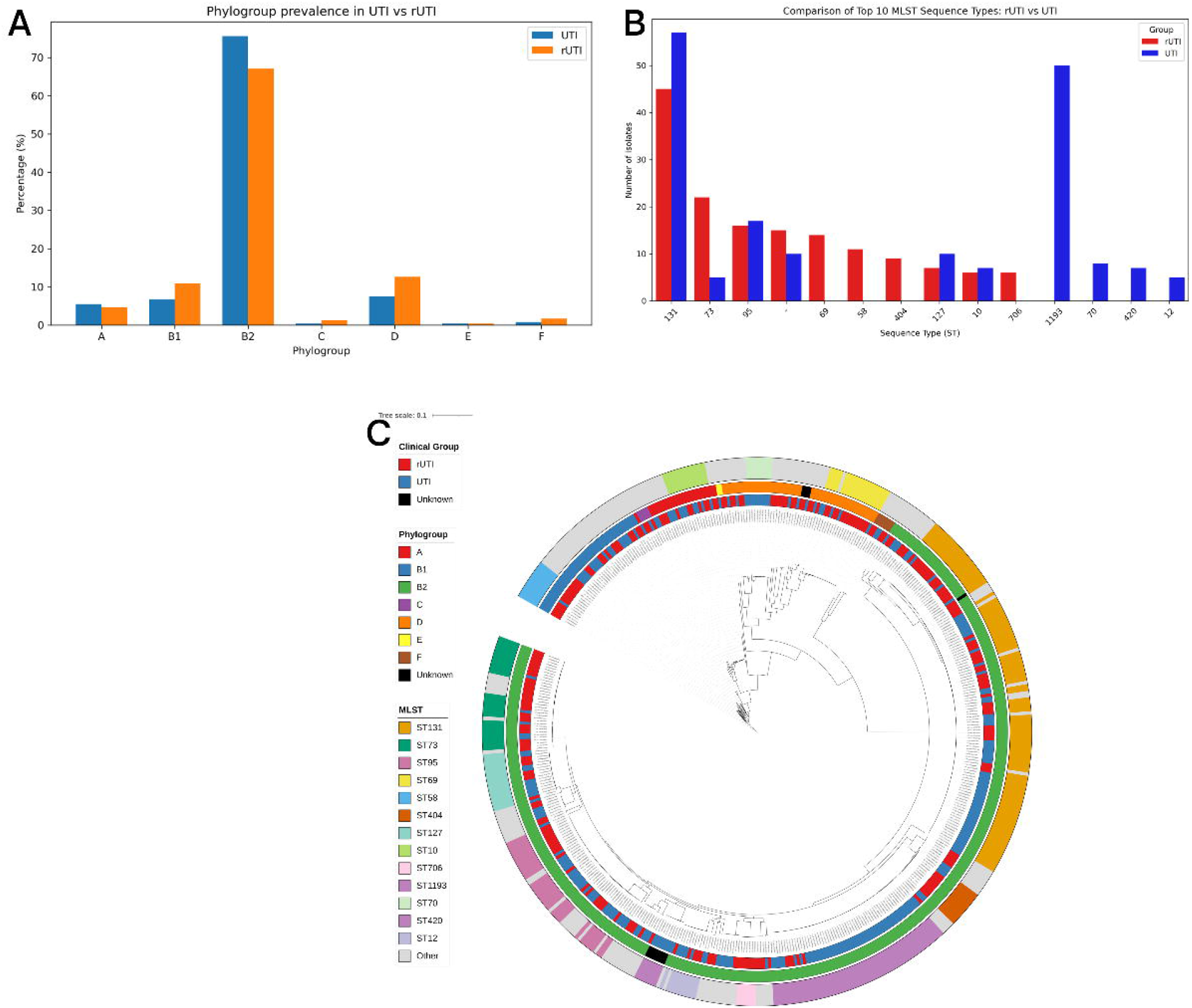

Collectively, all these findings suggested that both rUTI and UTI isolates showed comparable chromosomal diversity, similar pan-genome openness indicating equivalent rates of gene gain and loss with no distinct clades, clusters observed between the two phenotypes exhibit similar lineage distributions. Although several sequence types were observed exclusively or preferentially within one group (for example, ST69, ST58, and ST404 among rUTI isolates, and ST1193, ST70, ST420, and ST12 among sporadic UTI isolates), these lineages did not correspond to phenotype-specific clades in the recombination-aware core genome phylogeny (figure 3C). Instead, isolates belonging to both clinical phenotypes were broadly interspersed across the tree, with major ExPEC lineages such as ST131 containing representatives from both rUTI and sporadic UTI. This phylogenetic intermixing indicates that recurrent infection is not restricted to a specific evolutionary lineage. These findings robustly reject the hypothesis 1 that rUTI is solely associated with specific clonal expansions or distinct *E. coli* lineages. Instead, our results suggest that the recurrent phenotype is likely driven by accessory genomic determinants distributed across diverse genetic backgrounds. This realization necessitated a genome-wide association study (GWAS) to identify specific accessory genetic elements that may contribute to recurrent infection independent of chromosomal lineage.

### Enrichment of Conjugative plasmids in recurrent UTI

To support the hypothesis that rUTI-associated *E. coli* differs from sporadic UTI-associated isolates at the plasmidome level, we compared replicon composition, mobility potential, relaxase diversity and Mating pair formation system (MPF) between rUTI and UTI (figure 4). IncFIA, IncFIB were slightly predominant in UTI compared to rUTI representing the most frequently detected plasmid incompatibility groups. Additional replicon types, including IncFII, Col156 and Incl1/B/O were also detected at lower counts in both phenotypes where most of the replicons were slightly abundant in UTI isolates (figure 4A). Replicon analysis revealed that no unique replicon types associated with recurrence. Conjugative plasmids were more frequently observed in rUTI isolates whereas non-mobilizable and mobilizable plasmids were comparable between the two phenotypes (figure 4B). The MOBP relaxase family was enriched in rUTI and MOBQ family in UTI isolates. MOBF was consistently present in both phenotype groups (figure 4C). Other relaxase such as MOBH, MOBC, MOBV were present at lower counts (figure 4C). MPF_F was the most prevalent mating pair formation system in both the groups, however slightly enriched in rUTI. While MPF_I and MPF_T were found at lower counts in both the groups, most of the substantial proportion of plasmids couldn’t be assigned to a defined MPF category (figure 4D). Collectively plasmidome characterization revealed enrichment of conjugative plasmids and mobility associated features in rUTI, suggesting a potential role of plasmid-mediated horizontal gene transfer in recurrence. This supported the hypothesis 3 that rUTI-associated *E. coli* isolates exhibit consistent plasmid-level differences compared with UTI isolates.

**Fig 4.**
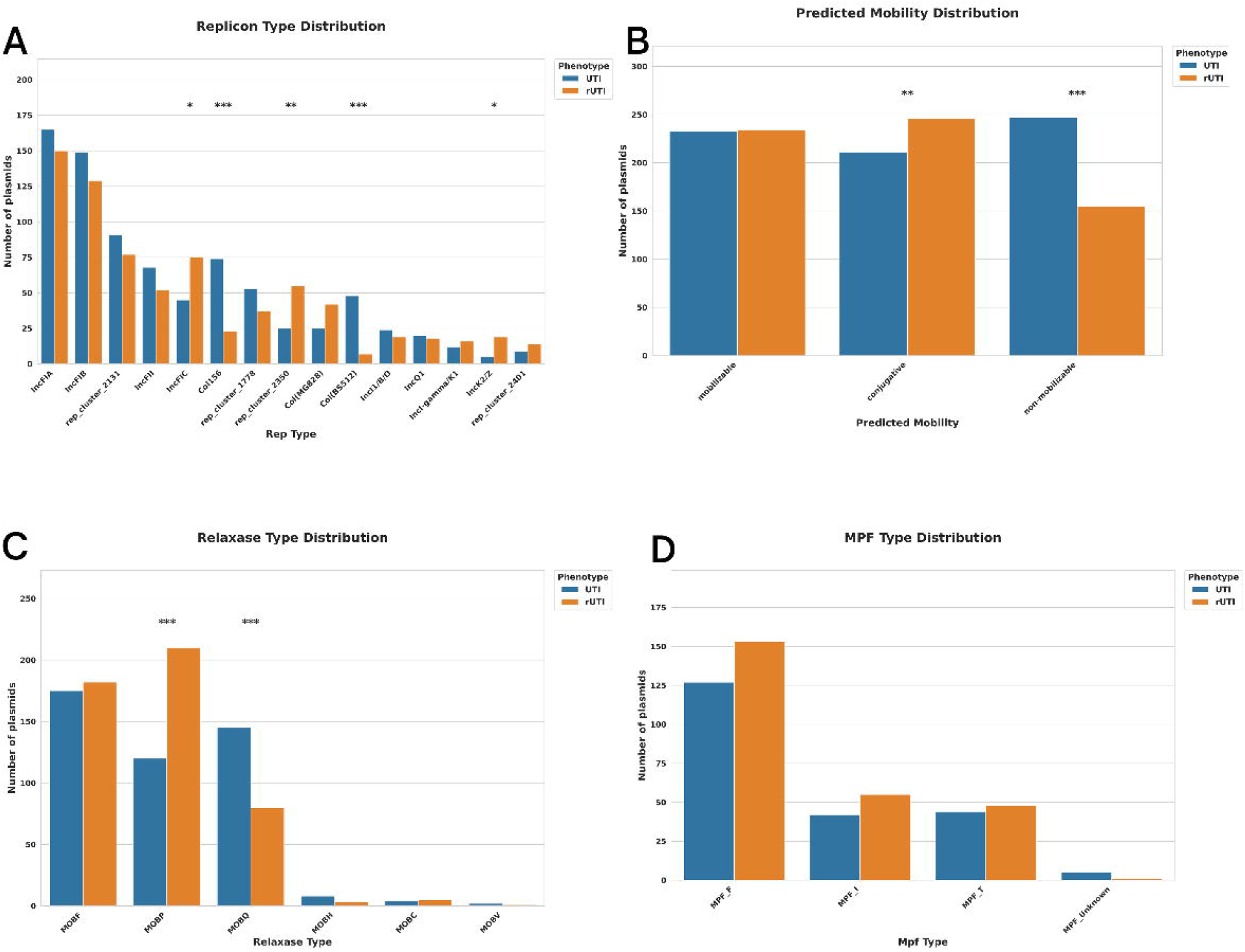

### Integrated chromosome and plasmid features enable machine-learning discrimination of rUTI isolates

To detect whether pathogen derived genomic features associated with rUTI are either encoded in chromosomal genes, plasmid-borne genes, or their combined genomic context, we performed 5 different machine learning models across these genomic compartments. Feature selection was performed using Chi-square, mutual information and Boruta selection independently for each genomic compartment datasets. As a result of Chi-Square testing feature selection, 2263 genes from chromosomal datasets, 456 genes from plasmids and 2655 genes from the combined datasets were selected as significant features (appendix 3 pp 2). For the chromosomal datasets mutual information selects 3986 genes, 1205 genes from plasmids and 5032 from combined datasets. Similarly using Boruta feature selections, 54 genes from chromosomal, 5 genes from plasmids and 78 genes from the combined datasets as significant features. Features from all the three methods named as STRICT and RELAXED Features (based on intersection and Union of all features from 3 different methods) and as a result for chromosomes 41 genes were resulted as STRICT and 4737 as RELAXED, 4 genes as STRICT and 1354 as RELAXED for plasmids and 56 as STRICT and 5906 as RELAXED features for combined datasets which were taken for further model building (appendix 5). Multiple supervised ML classifiers (XGBoost, Random Forest, ExtraTrees, Logistic Regression, and SVM) were trained under STRICT and RELAXED feature-selection regimes. Across models, plasmid-only feature sets yielded inconsistent predictive performance, with no single model or feature space emerging as dominant. In contrast, models built on the combined genome dataset achieved the highest overall test accuracy compared to chromosome-only models, particularly reaching ∼0.98 (figure 5A) in the best-performing SVM configuration and area under the ROC curve (figure 5B). In addition, the combined datasets ML model, constructed using either STRICT or RELAXED features, consistently displayed better accuracy in all the models; however, models constructed using STRICT features exhibited better performance with a limited number of features compared to models with a very large number of RELAXED features. This indicates that recurrence-associated genomic signals are present in both chromosomal and plasmid compartments, with increased discriminatory capacity when genomic compartments are integrated and isolating either compartment alone diminishes predictive power and thereby supports hypothesis 2.

**Fig 5.**
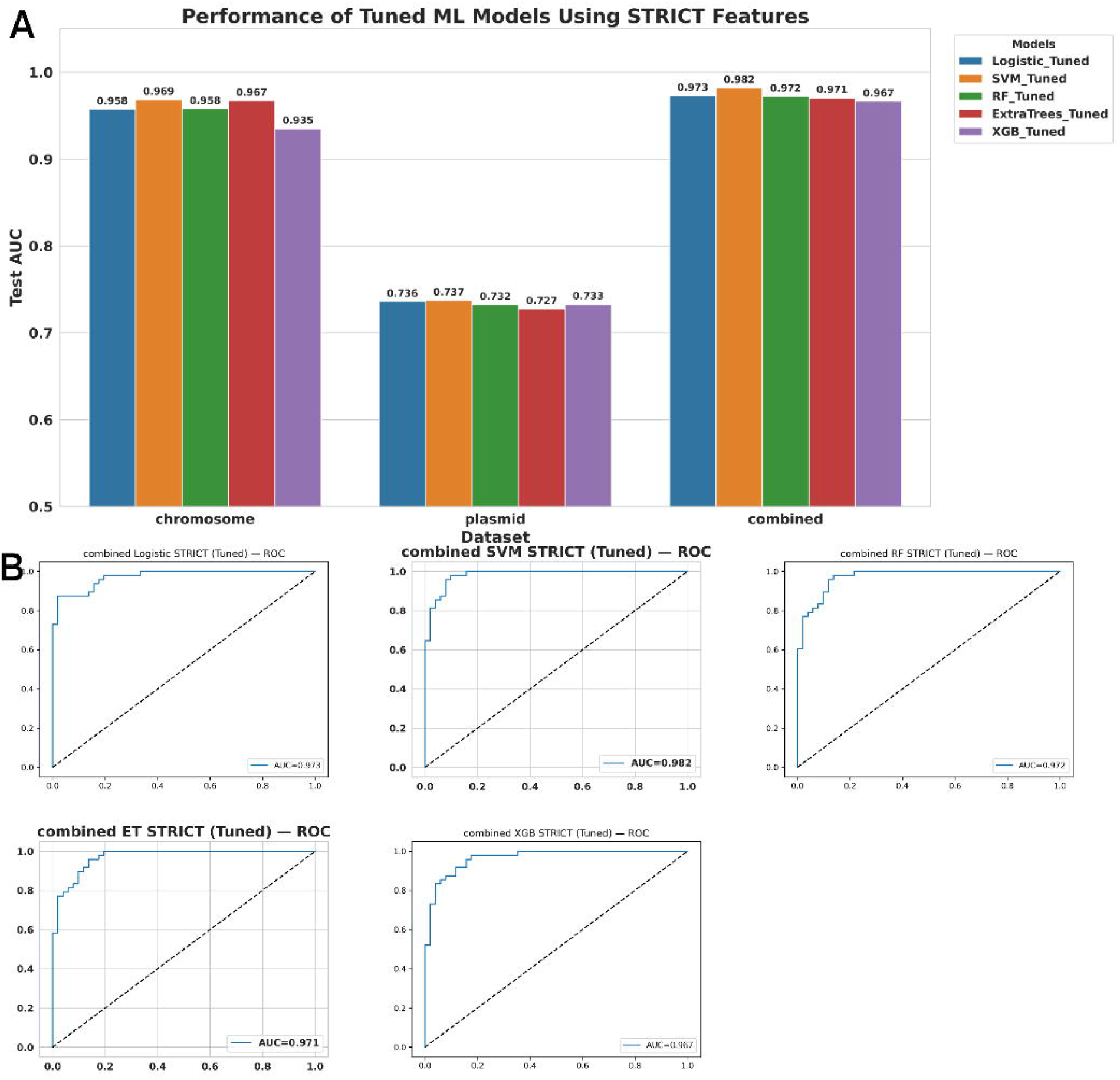

### Integrated ML-PanGWAS analysis reveals toxin-anti toxin systems and polygenic signal

To identify biologically meaningful genetic signals that could distinguish rUTI from sporadic UTI isolates, we integrated supervised machine learning with pan-genome-wide association analysis of features taken from the combined genomic datasets. To ensure robust detection of rUTI-associated genetic features, we applied two complementary feature discovery approaches in GWAS, Pyseer and Scoary (appendix 6). The latter evaluates associations between gene presence-absence and phenotype using contingency table-based statistics, whereas Pyseer applies a linear mixed model (LMM) incorporating a genetic relatedness (kinship) matrix to control for population structure. By accounting for shared ancestry among isolates, the Pyseer framework reduces false-positive associations attributable to clonal expansion or lineage effects. Pyseer identified 68 genes significantly associated with rUTI phenotype, in parallel Scoary detected 296 genes significantly associated with rUTI. Functional annotation of pyseer-associated genes revealed significant overrepresentation of biological processes related to transcriptional regulation, bacteriocin immunity, and sensory transduction (appendix 7). Similarly GO analysis of genes filtered using Scoary showed enrichment in biological categories such as host colonization, virulence, and horizontal gene transfer (appendix 7). Overlapping functional themes between Scoary and Pyseer support a distributed genomic architecture rather than a single dominant virulence determinant underlying rUTI. To further ensure the robustness of the detected genetic signals, we have taken the subset of genes that were consistently identified across pyseer, scoary and ML (appendix 6). Intersection of genes revealed that Pyseer ∩ Scoary: n = 30, Scoary ∩ ML: n = 16, Pyseer ∩ ML: n = 12 and Pyseer ∩ Scoary ∩ ML: n = 10 (figure 6a). Notably, 3 genes namely *cbeA, cbtA*, and *ldrD* followed by the (7 hypothetical genes/ clusters namely group_10205, group_10462, group_5473, group_6673, group_6803, group_8601, group_9085) were consistently detected across all three analytical frameworks (Pyseer ∩ Scoary ∩ ML), representing the most robust rUTI-associated genomic signals.

**Fig 6.**
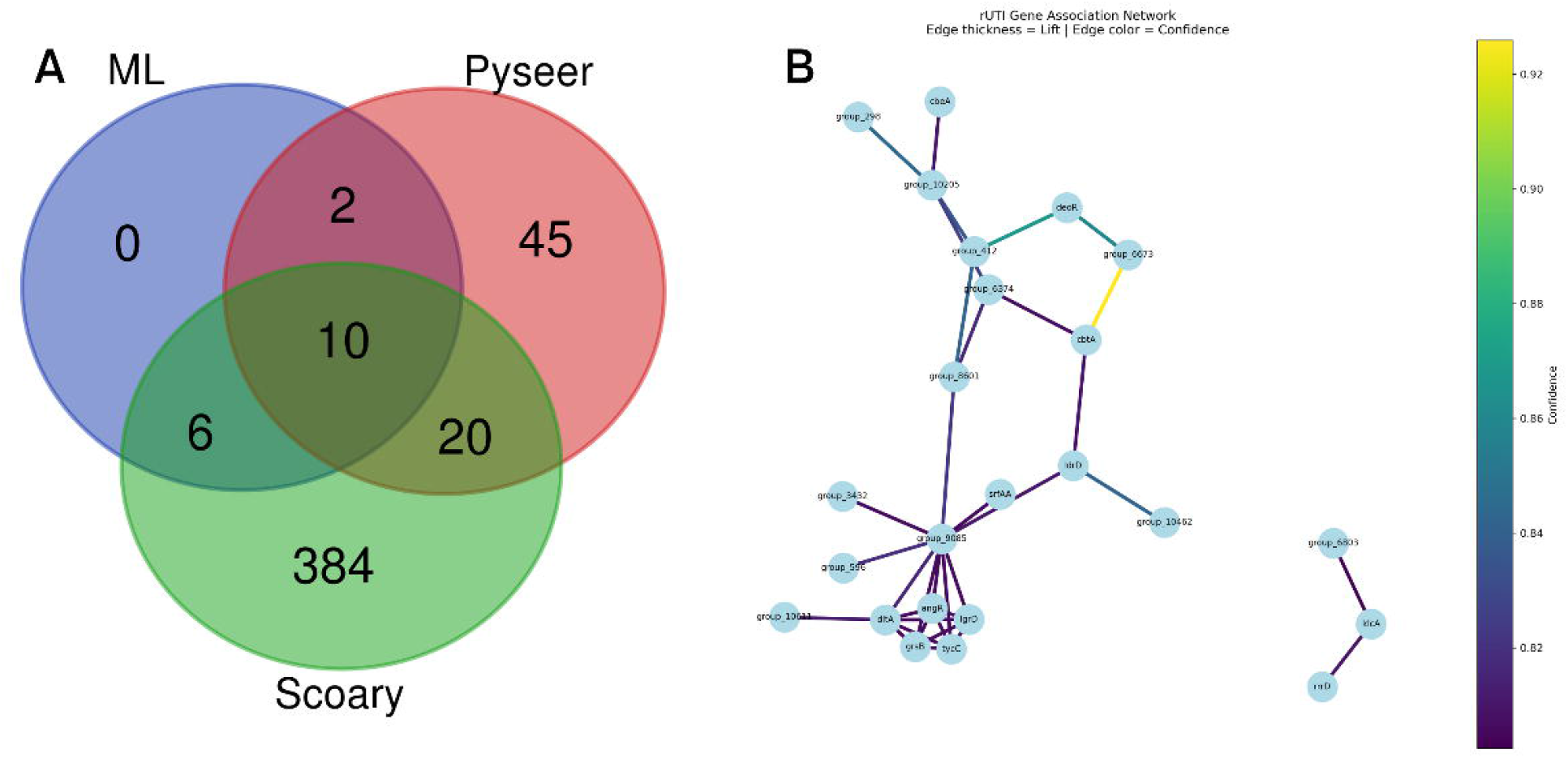

Together, these findings support hypothesis 4, suggesting that rUTI is driven by coordinated gene networks involving persistence, stress adaptation, and regulatory modulation, rather than by a single high-penetrance virulence factor. To move beyond the identification of statistically robust individual gene candidates, we applied seeded association rule mining using the Apriori algorithm to evaluate gene co-occurrence patterns within rUTI isolates. This approach enabled the identification of cooperative gene combinations enriched specifically in recurrent cases (appendix 6). A total of 23 significant rUTI specific rules in which (12 annotated genes and twelve hypothetical proteins, participated in high-confidence ranges from 0.80 to 0.93 association rules, with lift values exceeding 1.5, indicating non-random co-occurrence of gene combinations that were significantly enriched in rUTI isolates (Table 1). These rules revealed recurrent co-occurrence between toxin-antitoxin system components, metabolic regulators, and stress-response genes, indicating structured co-inheritance rather than random gene presence (figure 6b). Notably, several of the genes identified through integrated ML–PanGWAS analysis appeared within these high-confidence association rules, further supporting their role within cooperative genomic modules.

**Table 1.** Key recurrence-associated genes identified through association rule mining and their predicted functional roles in recurrent urinary tract infection.

| . S. No | Gene Name | Description | Possible role in rUTI |
| --- | --- | --- | --- |
| 1 | *cbtA* | Toxin of a type IV toxin-antitoxin system | Cytoskeletal proteins (FtsZ/MreB) (Target). Growth arrest/cell shape change. Enhances persistence, allows survival in antibiotic/host environment (Heller et al., 2017, Harms et al., 2018) |
| 2 | *deoR* | DeoR family transcriptional repressor of sugar metabolism | *DeoR* regulates nucleoside catabolism via repression of the deo operon, which may support metabolic adaptation under nutrient-limited conditions in the urinary tract (Engels & Wendisch, 2007) (Tourneux et al., 2000) |
| 3 | *ldrD* | Component of toxin-antitoxin (Type I) system | Translation blocking via antitoxin RNA. Growth arrest. Promotes stress survival and persistence (Harms et al., 2018) |
| 4 | *rrrD* | Encoded in the cryptic lambdoid prophage DLP12; essential for lysis of bacterial cell wall, exhibits peptidoglycan hydrolyzing activity; belongs to glycosyl hydrolase 24 family (165 aa). | Modulates biofilm formation by controlled cell wall remodeling; mutants show defective biofilm architecture, which can affect persistence and colonization in urinary tract infections (Toba et al., 2012). |
| 5 | *klcA* | Anti- restriction/plasmid- encoded protein | *KlcA* presence increases the transformation efficiency of plasmids in *E. coli* by ~4–6× when Type I RM systems are present.<br>This facilitates conjugative plasmid establishment, which often carry virulence or persistence genes, including those relevant to recurrent urinary tract infection (rUTI). (Nielsen et al., 2021) |
| 6 | *cbeA* | Toxin of a type IV toxin- antitoxin system | CbeA, the antitoxin component of the type IV *cbeA-cbtA* system, stabilizes cytoskeletal proteins (MreB and FtsZ) and counteracts toxin-induced growth inhibition (Singh et al., 2021; Nielsen et al., 2021) |
| 7 | *dltA* | Uroepithelial adhesion (LTA alanylation); direct mouse UTI model | the *dlt* operon is responsible for the D-alanylation of LTA and includes *dltA*, which encodes the D-alanyl carrier protein ligase which appears to limit UTI virulence because mutations in the *dlt* synthetic genes increase colonization of urothelial cells in vitro and in vivo, suggesting that their expression interferes with the host-pathogen interaction in the urinary tract (Wobser et al., 2014) |
| 8 | *higB* | Toxin of a type II toxin- antitoxin system | mRNA cleavage → translation inhibition<br>And there by could relate to dormancy/persister formation (Harms et al., 2018; Wang and Wood., 2011, Shah et al., 2006) |
| 9 | *angR* | Siderophore biosynthesis regulator (iron acquisition) | *angR* is a bifunctional regulatory/biosynthetic gene of siderophore. it likely represents a siderophore regulatory component of a pathogenicity island co-located with the other NRPS genes (*grsB*, *lgrD*, *tycC*) in this cluster (Mike et al., |
| 10 | *grsB_1* /<br>*lgrD* /<br>*tycC* — | NRPS (Non-Ribosomal Peptide Synthetase) modules (Secondary metabolic islands) | 2018; Henderson et al., 2009)<br>NRPS-associated genomic islands have been reported in pathogenic *E. coli* lineages, including UPEC, and may indirectly support persistence or colonization within the urinary tract (Mike et al., 2018) |

### External Validation of the ML Model

To assess the generalisability of the combined-genome Logistic Regression model, an independent external dataset comprising 63 *E. coli* genomes (31 rUTI, 32 sporadic UTI) was used for validation. The model trained on the primary cohort of 491 isolates was applied directly to the external cohort using the same STRICT feature space derived from the combined chromosomal and plasmid pan-genome. The external validation yielded an overall accuracy of 70.0%, substantially lower than the internal test accuracy of ∼98%. This performance gap is consistent with known challenges in cross-cohort transfer of pan-GWAS-based predictive models and reflects expected heterogeneity between training and external datasets, including differences in geographic origin, sequencing platforms, assembly quality distributions, and clinical phenotyping methodology.

## Discussion

Recurrent urinary tract infections (rUTI) remain major clinical challenge where several studies reported pathophysiology in terms of bacterial persistence with intracellular bacterial communities (IBCs), quiescent intracellular reservoirs, biofilm formation, reinfection from intestinal reservoirs driven by incomplete antibiotic therapy. However, the underlying genomic architectures that distinguish rUTI from sporadic UTI remains poorly understood. Most of the studies focused on virulence genes, antimicrobial susceptibility, phylogenetic backgrounds yet recurrence does not correlate with specific clonal lineages or individual virulence genes. To address this gap, the current study employed ML integrated PanGWAS combining lineage analysis, plasmidome characterization to determine whether rUTI is driven by distinct clonal expansion or by genomic configurations that is distributed across chromosomes and plasmids. By analysing the recombination aware phylogeny, phylogroup distribution, core genome PCA, nucleotide diversity, genetic shrinkage and pangenome openness confirmed that rUTI is not lineage dependent since there is no distinct clustering that separates rUTI from UTI. This aligns with findings that lineage or clonal expansion alone could not explain recurrence (Nielsen et al., 2021). While phylogroups D showed slight enrichment in rUTI, consistent with prior literature reported for the ability of these groups as reservoirs of drug-resistant genes (Whelan et al., 2023; Mendonça et al., 2011). Isolates responsible for persistence or relapse have also been frequently linked to phylogroup D backgrounds. Notably, sequence type ST69, belonging to phylogroup D, was frequently represented in rUTI isolates, underscoring the correlation between antimicrobial resistance and recurrence (Hidad et al., 2022). These observations support the notion that certain non-dominant phylogenetic groups may disproportionately contribute to recurrence-associated traits, including biofilm formation, antibiotic tolerance, and persistence. In alignment with the recent study our observations indicate despite being numerically smaller than B2 and D, phylogroup B1 demonstrates a notable increase in prevalence with rUTI than in UTI cases (Mahshouri et al., 2025). Our findings support the concept that commensal *E. coli* belonging to phylogroups A and B1 (Dadi et al., 2020; Yazdanpour et al., 2020), may contribute to recurrence, as these lineages can act as the reservoirs of UPEC associated virulence determinants, including adhesion genes such as *fimH* likely through accessory genetic elements (Ketkhao et al., 2024). Notably, several globally distributed extraintestinal *E. coli* lineages, including ST69, ST58, ST404 and ST73, were either uniquely detected (ST69, ST58, ST404) or enriched (ST73) among rUTI isolates in our study. These sequence types are all recognised UPEC or emerging ExPEC clones with documented associations with urinary tract infection, multidrug resistance and/or long□term host colonisation in previous clinical and genomic studies. Rather than defining a single “rUTI clone”, their distribution in our phylogeny supports a model in which recurrent disease arises when particular high□risk lineages acquire specific combinations of plasmid□borne maintenance systems, toxin– antitoxin modules and metabolic adaptors, as captured by our integrated ML–PanGWAS approach, thereby reinforcing that recurrence is a polygenic background□dependent phenotype rather than a property of one sequence type alone (figure 7). In plasmidome analysis, enrichment of conjugative plasmids indicates enhanced horizontal gene transfer potential among rUTI strains compared to sporadic isolates. The higher prevalence of MOBP, alongside IncF replicons and MPF_F systems, suggests that rUTI-associated *E. coli* utilize a highly efficient ‘mating’ machinery. Similarly, there is a significant enrichment of MOBP relaxase types, capable of transfer across diverse gram-negative bacteria (Garcillan-Barcia et al., 2009).

**Fig 7.**
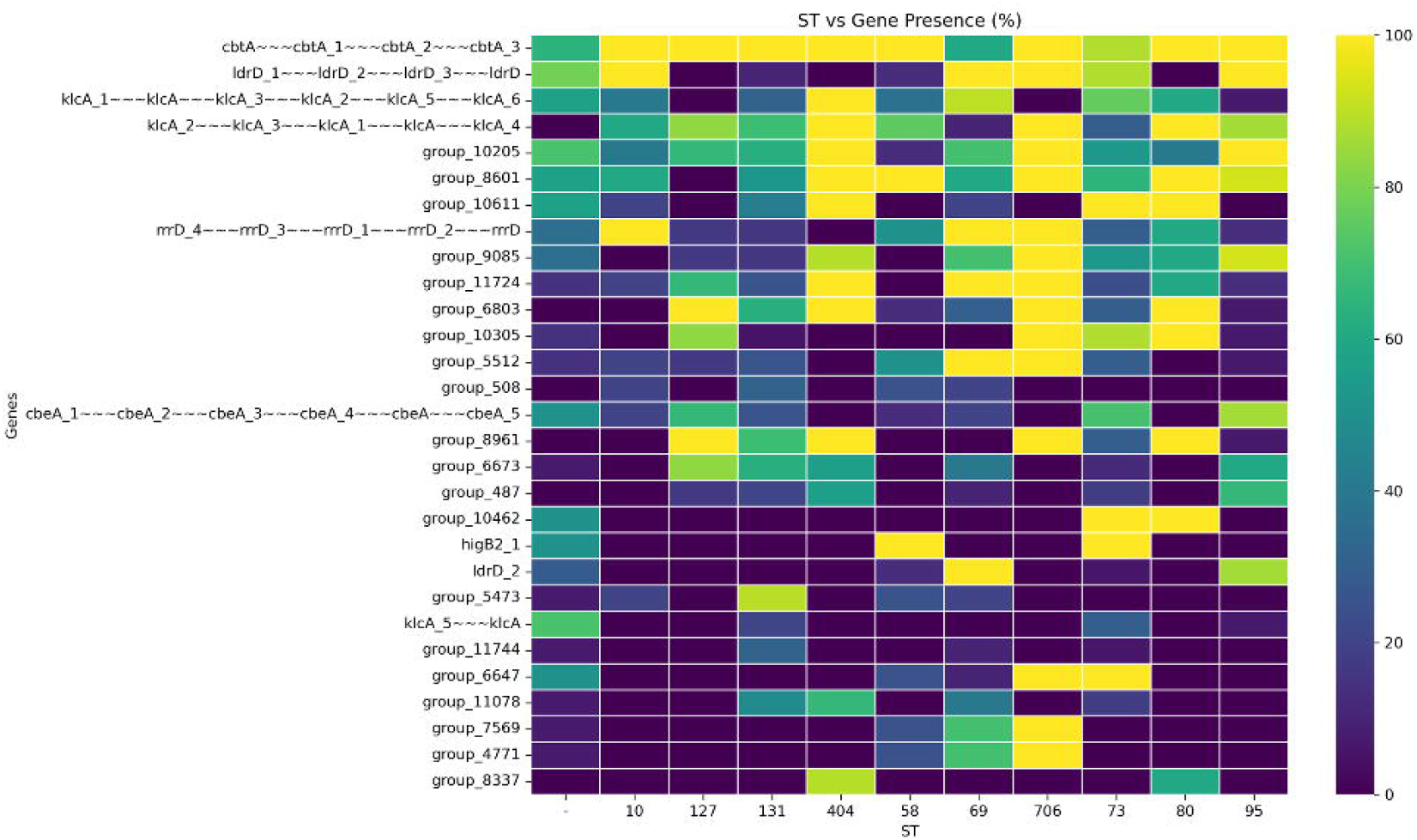

Feature selection and ML modelling demonstrated that rUTI-associated signals are highly distributed across the genome. Although thousands of features showed statistical association (Chi^2^ and mutual information), only a small subset remained robust after Boruta filtering, and predictive performance improved when features were integrated. This pattern indicates that recurrence is not driven by a single dominant virulence determinant but rather by polygenic, weakly penetrant signals. Similar distributed genomic architectures have been reported in bacterial GWAS studies of virulence and persistence, where combinations of accessory genes, metabolic loci, and stress-response determinants collectively influence phenotype (Lees et al., 2018).

The modest effect sizes of individual genes but improved performance in integrated models strongly support a cooperative gene network model of recurrence. A striking result of the integrated ML-PanGWAS analysis was the convergence of toxin-antitoxin (TA) system components, including: *cbtA, ldrD, cbeA* and *rrrD*. TA systems are well-established mediators of bacterial persistence. Type II toxins such as *CbtA* inhibit cytoskeletal polymerization and cell division, inducing growth arrest (Heller et al., 2017). *HigB* toxins inhibit translation by mRNA cleavage, promoting dormancy and antibiotic tolerance (Schureck et al., 2015). *LdrD* belongs to type I TA systems that can induce membrane damage and growth inhibition. TA systems contribute to the formation of persister cells subpopulations that survive antibiotic treatment without genetic resistance (Harms et al., 2016). Persistent intracellular UPEC reservoirs in bladder epithelial cells are a major mechanism underlying recurrence (Hannan et al., 2010). Thus, enrichment of TA modules provides a mechanistic explanation linking genomic architecture to clinical recurrence. Beyond TA systems, recurrence-associated genes included *deoR* (transcriptional regulator of nucleotide metabolism), *gntK* (gluconokinase involved in gluconate metabolism), *cbeA* (nutrient uptake associated) revealed that metabolic flexibility is increasingly recognized as central to UPEC persistence. Utilization of alternative carbon sources such as fucose and gluconate enhances survival within the nutrient-limited urinary tract (Alteri et al., 2009). Fucose metabolism has been implicated in intestinal colonization and niche adaptation, potentially facilitating gut-bladder transmission cycles that contribute to recurrence. DeoR-mediated regulation of nucleotide metabolism may influence stress survival and growth modulation, linking metabolic control to persistence strategies. These findings align with the concept that metabolic rewiring complements virulence in sustaining chronic recurrent infection. The convergence of lineage-independent distribution, plasmid enrichment, persistence-associated TA systems, and metabolic adaptation genes supports a revised model of recurrence in rUTI which is driven by cooperative gene networks involving stress tolerance, dormancy induction, metabolic flexibility, and plasmid stability rather than by single classical virulence genes. This model explains why traditional virulence profiling often fails to distinguish recurrent from sporadic UTI isolates. Instead, recurrence appears to emerge from subtle but coordinated genomic configurations that enhance survival under antibiotic pressure and host immune stress.

Several limitations of this study warrant consideration. First, the construction of the sporadic UTI control group from publicly available genomes introduces inherent phenotypic label noise: isolates classified as sporadic UTI were labelled based on their status at the time of sequencing, without long-term follow-up to confirm the sustained absence of recurrence. Since approximately 25-30% of women experience at least one recurrent episode following a first UTI, a meaningful fraction of sporadic control genomes may represent undetected recurrent cases. This blurring of the true phenotypic boundary is an unavoidable constraint of all pan-GWAS studies relying on public genomic databases and is expected to attenuate both internal model accuracy and external transferability. The observed gap between internal (∼98%) and external (70%) validation accuracy is consistent with this interpretation, reflecting both phenotypic label noise and dataset heterogeneity rather than model overfitting alone. Second, the sample size, while among the largest assembled for rUTI pan-GWAS to date, may have insufficient power to detect small-effect accessory gene associations. Third, the absence of experimental validation means that putative functional roles of identified genes particularly the hypothetical proteins in the three-way intersection remain to be confirmed. Future studies combining prospective cohort design, standardised phenotypic classification, and functional genomics will be essential to address these limitations.

## Conclusion

This study provides a comprehensive genomic investigation of recurrent urinary tract infection (rUTI)-associated *E. coli* using an integrated framework combining pan-genomics, plasmidome characterization, machine learning and genome-wide association analysis. Comparative population analysis demonstrated that rUTI isolates do not form distinct phylogenetic lineages and share similar chromosomal diversity and open pan-genome architecture with sporadic UTI isolates, indicating that recurrence is not driven by clonal expansion. Instead, plasmidome analysis revealed subtle enrichment of conjugative plasmids and mobilization systems in rUTI isolates, suggesting an increased potential for horizontal gene transfer. Machine learning models further showed that integrating chromosomal and plasmid features significantly improves the discrimination of rUTI isolates, supporting a distributed genomic signal underlying recurrence. Integration of ML feature selection with Pan-GWAS converged on toxin–antitoxin systems, metabolic regulators and stress-response genes, while association rule mining revealed cooperative gene modules enriched in rUTI isolates. Collectively, these findings support a model in which recurrent infection arises from coordinated genomic configurations involving persistence mechanisms, metabolic flexibility and plasmid-mediated gene exchange rather than from single virulence determinants. This integrative genomic framework provides new insights into the evolutionary and functional basis of rUTI and highlights potential molecular targets for improved surveillance and therapeutic intervention.

## Data Availability

All genome data analysed in this study are publicly available from the National Centre for Biotechnology Information (NCBI) Assembly database and the European Nucleotide Archive (ENA). Accession numbers and associated metadata for all isolates included in this study are provided in Supplementary Data 1. Curated phenotype annotations (recurrent vs sporadic UTI), processed gene presence-absence matrices, and analysis scripts will be made available in a public repository (GitHub; link to be provided upon publication).

## Author Contributions

SM and SN conceived and designed the study. SR performed data curation, formal analysis, and interpretation of the data. SR drafted the manuscript. SM and SN contributed to critical revision of the manuscript for important intellectual content. All authors had full access to all the data in the study and approved the final version for submission.

## Declaration of interests

No competing interests were disclosed.

## Declaration of generative AI and AI-assisted technologies in the writing process

During the preparation of this work, the authors used ChatGPT-4 in order to improve readability. After using this tool, the authors reviewed and edited the content as needed, and take full responsibility for the content of the published article.

## Acknowledgments

The authors acknowledge the Department of Biotechnology (DBT), Government of India (grant number: BT/PR40150/BTIS/137/81/2023) and the SASTRA TRR grant (SASTRA TRR SCBT OCT-23) for providing computational resources at DBT-Bioinformatics Center at SASTRA Deemed to be University for performing the analysis.

## Notes

### Competing Interest Statement

The authors have declared no competing interest.

