## appendix 3 for "Integrated Machine Learning-PanGWAS Reveals Chromosome-Encoded Persistence Networks and Plasmid Plasticity in Recurrent Urinary Tract Infection in *Escherichia coli*"

Supplementary File

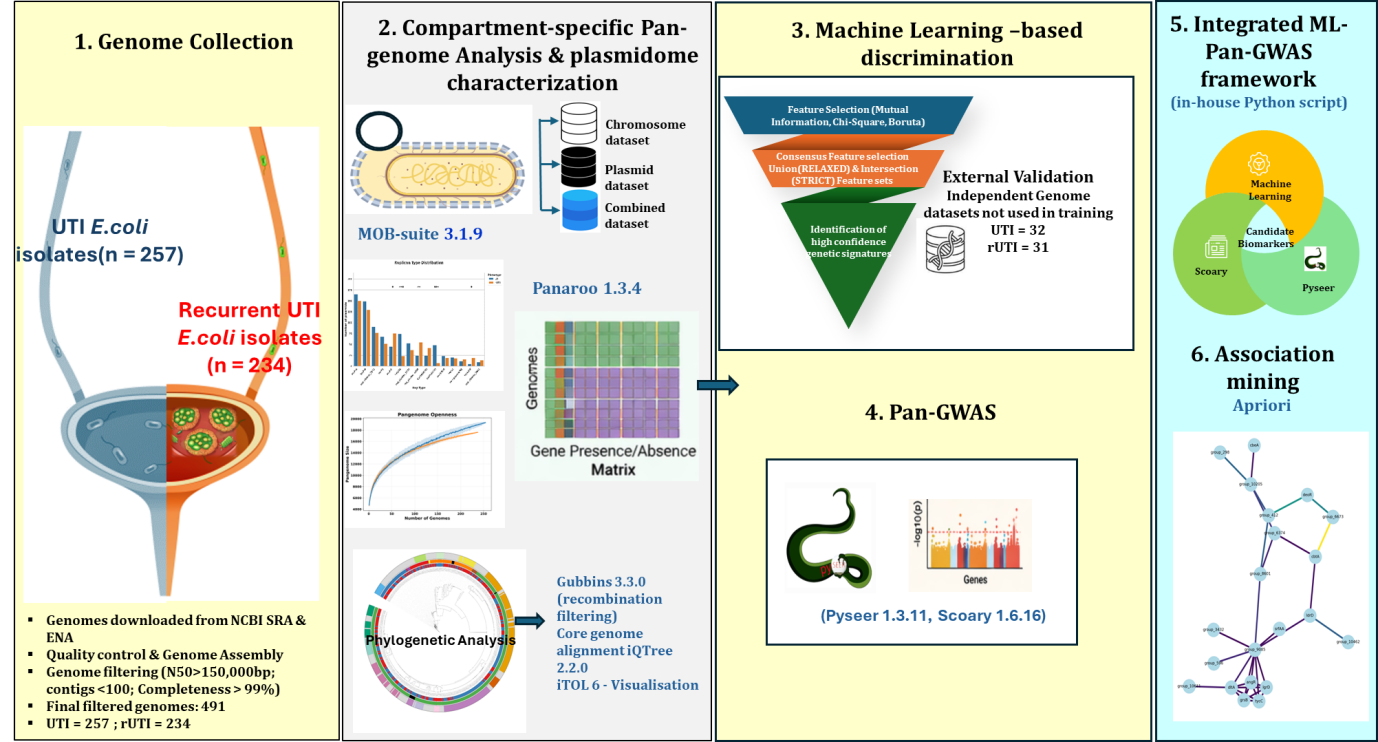

Fig S1. Workflow for the integrated Pan-GWAS and Machine Learning framework to discriminate rUTI and UTI isolates

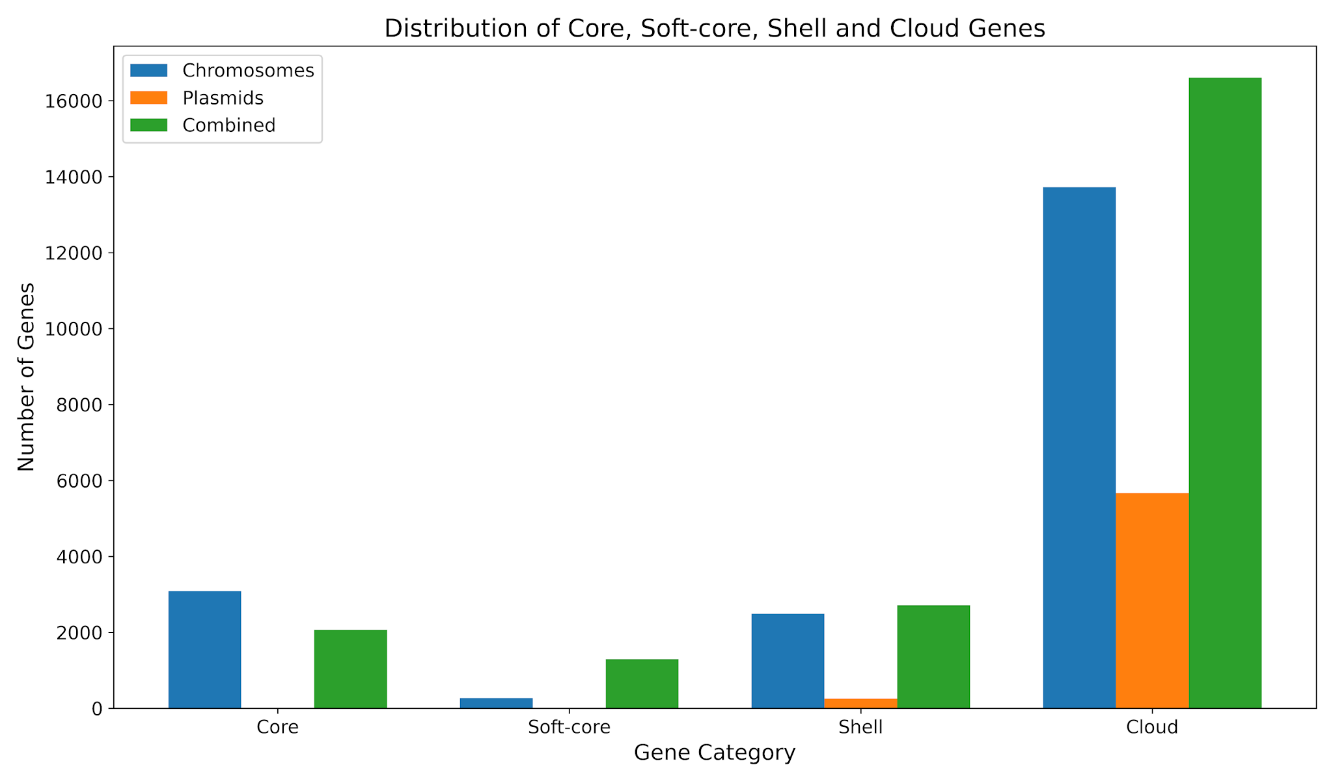

Fig.S2. Distribution of Core, Soft-core, Shell and Cloud genes among *E.coli* isolates across  genomic dissection.
